# Opportunities to improve nutrition delivery in hospital after discharge from an intensive care unit: A mixed methods analysis

**DOI:** 10.1101/2023.03.31.23288012

**Authors:** Sarah Vollam, Owen Gustafson, Lauren Morgan, Natalie Pattison, Oliver Redfern, Hilary Thomas, Peter Watkinson

## Abstract

**Background and Aims:** Though adequate nutrition following critical illness is fundamental to rehabilitation, it is poorly provided. To inform interventions to improve nutrition support for patients discharged from an intensive care unit (ICU), we aimed to document remediable problems in nutrition management on general hospital wards, and the context for these problems.

**Methods:** This work forms part of a larger mixed methods study: REcovery FoLlowing intensivE Care Treatment (REFLECT). From three NHS hospitals, chosen to represent different hospital settings, we conducted in-depth reviews of 20 cases where in-hospital death after ICU discharge was judged ‘probably avoidable’ and 20 cases where patients survived to hospital discharge. We interviewed 55 patients, family members and staff about their experiences of post-ICU ward care. From these primary data we extracted information related to nutrition provision to develop a process map of how enteral feeding is delivered to patients on hospital wards after ICU discharge.

**Results:** Problems with nutrition delivery were common (81 problems in 20/40 cases), mostly (70/81) in patients whose death was judged “probably avoidable”. Common issues included failure to monitor nutritional intake, delays in dietician/nutritional support referrals, removal of enteral feeding tubes before oral intake was established, and poor management of enteral nutrition delivery. Staff identified workload related to the high care needs of post-ICU patients as contributing to these problems in nutrition delivery. The process map of enteral feeding delivery demonstrated that local policy for tube placement confirmation risked prolonged system-related delays to administering naso-gastric feed, significantly affecting the volume of feed delivered to patients.

**Conclusions:** Using a novel mixed methods approach, we identified problems throughout the process of delivering nutritional support, which had profound consequences for post-ICU patients. We demonstrated the importance of multi-professional collaboration in delivering enteral nutrition. Improving collaborative working processes within the ward system may ensure timely confirmation of correct nasogastric tube placement, and support safe feeding. Addressing the common problems in post-ICU nutritional support we identified may support improved nutritional delivery and potentially enhance recovery from critical illness.

**Study registration:** ISRCTN:14658054

## INTRODUCTION

Poor physical recovery following critical illness is common, affecting up to 25% of patients, especially the elderly and frail [1,2]. These effects may persist for years, and are associated with poor quality of life and depression [3,4]. This poor physical function is commonly attributed to muscle loss due to catabolism during critical illness [5]. Adequate nutrition is essential for regaining muscle mass, without which, patients are unlikely to maximise strength and mobility following critical illness [6]. The European Society of Parenteral and Enteral Nutrition (ESPEN) describe three metabolic phases of critical illness [7]. The two ‘acute’ phases – early and late – are characterised by catabolic stress response. During catabolism, nutrition cannot be utilised and muscle loss is likely, even when receiving enteral feeding [5]. During the post-acute third phase, anabolism may return. It is only once anabolism is reinstated that patients can utilise nutrition to support their recovery. ESPEN nutritional guidelines suggest the catabolic acute phases may last up to seven days [7]. Although very little evidence exists to support this, one study did demonstrate rapid recovery of gastric emptying in critically ill patients discharged from ICU, a sign of returning gastric function [8]. Therefore, for most patients, it is likely that this post-acute anabolic third phase will occur once they have been discharged from ICU to the ward, making this a crucial period for optimising nutrition, either orally or enterally, to support physical rehabilitation [9].

Much research into nutrition in critical illness focuses on the acute phase in ICU, missing the opportunity to maximise rehabilitation in this patient group. Three recent reviews identified that nutritional needs were often not met in the post-ICU in-hospital period, but there is little research into nutrition beyond the first seven days of critical illness [9–11]. There is some evidence that patients discharged from ICU fail to meet 50% of their nutritional needs through oral intake due to poor appetite and physical dependency making eating difficult [12,13]. [13]More information is needed to inform improvements in nutrition delivery during the recovery period following critical illness.

We conducted a mixed methods study of post-ICU ward care (REFLECT: REcovery FoLlowing intensivE Care Treatment), aiming to identify potential improvements to ward-based care provision for critical care survivors to improve outcome. In this paper, we aim to examine the problems in care related to nutrition delivery (including oral intake, enteral feeding and parenteral nutrition) and map the process of delivering enteral nutrition on the ward.

## MATERIALS AND METHODS

We report our study using a combination of the Strengthening the Reporting of Observational Studies in Epidemiology (STROBE) [14] for cohort studies, and Consolidated criteria for reporting qualitative research (COREQ) [15] reporting guidelines, in line with our mixed methods approach.

### Definitions

We defined nutritional support as provision of total parenteral nutrition; enteral nutrition; or help with oral feeding, including practical help and nutrition supplements. We also included fluid intake, as nutrition and hydration are intrinsically linked.

We defined ‘probably avoidable deaths’ according to published guidance on structured judgement reviews [16].

### Primary data collection

We collected data at three NHS hospitals, selected to represent different sized ICUs and provision of post-ICU services. We published the protocol [17], and registered the study prospectively: ISRCTN:14658054. Ethical approval was granted by Wales REC 4 (reference 17/WA/0139).

As provision of post-ICU ward care is complex and multifactorial, we undertook a mixed methods project using three approaches to examine care delivery, with each approach offering different contextual information about the problem. Through our previously published Structured Judgement Review of 300 patients who died on the wards following discharge from ICU at the three sites we identified 20 cases where death was probably avoidable [18]. In this paper we present nutritional information extracted from these cases, alongside an equal number of survivor cases from across the three sites. We also report data related to nutritional support from qualitative interviews with patients, family members and staff. We have taken this approach with other areas of post-ICU care delivery we identified as problematic: out-of-hours discharge from ICU [19] and mobilisation [20]. Methods are presented in brief below, with further details provided in our published protocol [17], completed STROBE and COREC checklists [14,15] and in supplementary material.

### Structured judgement reviews

To provide an overarching view of post-ICU care, we reviewed the medical records of 300 patients who were discharged from ICU and subsequently died in hospital, using an established mortality review method. The Structured Judgement Review (SJR) guides development of a brief narrative of the trajectory of care for each case, which contributes to an overall assessment of the avoidability of each death from ‘definitely not avoidable’ to ‘definitely avoidable’. This approach identified the 20 probably avoidable deaths. This work has previously been reported [18].

### In-depth reviews

To develop our understanding of the context of these cases, for each of the 20 probably avoidable deaths we used an established in-depth review framework methodology to guide identification of the problems in care for each case [21]. We report the problems related to nutrition and hydration here. However, the codes within the framework related to nutrition support were limited, with only two codes directly related to nutrition (3.10: other [nutrition] and 3.23: other [dehydration/malnutrition]). Other codes indirectly related to aspects of poor nutrition management, such as 3.16: inadequate handover and 3.17: lack of liaison with other staff, but these were limited and did not offer a clear account of the nutrition-related problems in care we identified. Therefore, for clarity, we have further categorised nutritional problems using codes derived from the narrative which forms part of each SJR.

For each of the problems in care identified we also selected a ‘contributory human factor’, from the established frameworks [21]. This promoted further consideration of the context of care documented in the care record for each patient, and allowed identification of the underlying reasons why problems may have occurred.

We also reviewed the records of 20 survivors across the three sites. Participants were a convenience sample, recruited through the same approach as the interviews (as describe in the protocol and supplementary information [17].

We drew anonymised vignettes from the in-depth reviews to illustrate common problems with nutritional support for patients discharged from ICU to the ward.

### Qualitative interviews

To broaden our understanding of the context of delivering care to this patient group, we conducted semi-structured interviews with 56 purposively sampled patients (n=18), family members (n=8) and staff (n=30) about their experiences of post-ICU ward care. We analysed these interviews using Braun and Clark’s six steps of thematic analysis [22]. Qualitative data related to nutritional delivery are reported in this paper. Further details about the approach taken are included in the published protocol and supplementary material [17].

### Functional Resonance Analysis Method

To develop our understanding of why nutritional problems in care occurred in this population, we used the primary data from the REFLECT study to map the delivery of nutrition to post-ICU patients on the ward, using the Functional Analysis Resonance Method (FRAM) [23]. This Human Factors (HF)-based approach aims to describe an ‘ideal world’ process to deliver the desired end goal – in this case successful delivery of nutrition to post-ICU patients on the ward - and the circumstances which contribute to this process. We identified specific activities within the process, integral to the delivery of the desired end goal (termed “functions”), and the circumstances needed to deliver each function (termed “conditions”). These conditions are split into: inputs, preconditions, resources, time constraints, and controls (Table 1). Two members of the study team (a nurse and a physiotherapist with both critical care and ward experience) with knowledge of the primary data from the REFLECT study developed the post-ICU nutrition FRAM. To ensure we did not miss any functions or conditions of the process not captured in the REFLECT data, we also invited a Critical Care Dietician with ward experience with experience at one site, and Medical Specialist Registrar with ward and ICU experience at two of the sites to help us develop the FRAM.

**Table 1.** Definitions and examples of FRAM conditions

| **FRAM Conditions** | **Definition** | **Example** |
| --- | --- | --- |
| **Function** | Activity in a process | Nurse commences NG feed |
| **Input** | Starts the function | Patient identified as needing NG feeding |
| **Precondition** | Must be satisfied before the function can start | NG aspirate confirmed at gastric contents through pH check |
| **Resource** | Needed to carry out function | NG feed pump |
| **Control** | Monitors or controls the function | Local NG guidelines |
| **Time** | Any time constraint that affects the function | Nurse time to administer feed |
| **Output** | The outcome of the function | NG feed commenced |

*Adapted from Clay-Williams et al. 2015 [24]*

We presented primary data to the site stakeholders. We then commenced the FRAM process with identification of the first function in the process of delivering nutrition to post-ICU patients on the ward, which was written on a coloured post-it note. We then discussed this function, prompting identification of all conditions of this function (preconditions, resources, etc.). We used different coloured post-it notes for each FRAM condition (input, precondition, etc.), and positioned these around the function. We repeated this process until it was agreed that we had identified all relevant functions associated with delivering nutrition to post-ICU patients on the ward. We then transcribed the finalised paper-based FRAM into the FRAM visualiser software (Version 0.4.1, May 2016: http://functionalresonance.com/FMV/index.html). The finalised FRAM is extensive, covering a wide range of functions related to nutrition delivery via various routes. For the purpose of this paper, we have limited the activities reported to those related to providing enteral nutrition, as this was an area identified as particularly problematic in the in-depth reviews. The full FRAM is included in supplementary material for reference.

## RESULTS

### In-depth reviews

We conducted 40 in-depth reviews (of 20 probably avoidable deaths and 20 survivors). Characteristics of these patients are provided in supplemental material table 1. During in-depth reviews, we identified problems with nutrition and fluid management in 20 of the 40 cases where we undertook in-depth review (table 2). Problems were more common in non-survivors than survivors (70 identified problems in 15 non-survivors versus 11 identified problems in 5 survivors, table 2). The coded problems occurred throughout the ward stay, and fell into three distinct categories of care delivery – handover between settings; monitoring and escalation of poor intake; and provision of nutritional support. We refer to the vignettes in table 3 to illustrate these problems.

**Table 2.** Nutritional problems in care delivery for non-survivors and survivors

| **Problem in care category**<br>*Frequency (n)* | **Non-survivors**<br>**(n=15)** | **Survivors**<br>**(n=5)** | **Total**<br>**n=20** |
| --- | --- | --- | --- |
| **Handover** |  |  |  |
| ICU handover/optimisation (nutritional and fluid) | 6 | 0 | 6 |
| **Nutritional Monitoring and Escalation** |  |  |  |
| Failure to recognise and act on poor intake | 5 | 1 | 6 |
| Dehydration/fluid management | 6 | 0 | 6 |
| Failure to monitor nutrition/fluid balance | 7 | 3 | 10 |
| Specialist referral not made/delayed/advice not followed | 10 | 0 | 10 |
| Blood result surveillance/supplementation | 7 | 1 | 8 |
| Recognising clinical problem related to nutrition | 4 | 1 | 5 |
| Acting on clinical problem related to nutrition | 4 | 2 | 6 |
| Escalating clinical problem related to nutrition | 4 | 0 | 4 |
| Delay in radiological investigation/review (diagnostic) | 4 | 0 | 4 |
| **Nutritional Support Provision** |  |  |  |
| Early cessation of enteral or total parenteral nutrition | 3 | 0 | 3 |
| Ensuring secure NG placement | 2 | 2 | 4 |
| Delay in radiological investigation/review (related to NG placement) | 1 | 1 | 2 |
| Escalation of nutritional support when route failed | 4 | 0 | 4 |
| Team leadership | 3 | 0 | 3 |
| Total | 70 | 11 | 81 |

**Table 3:**
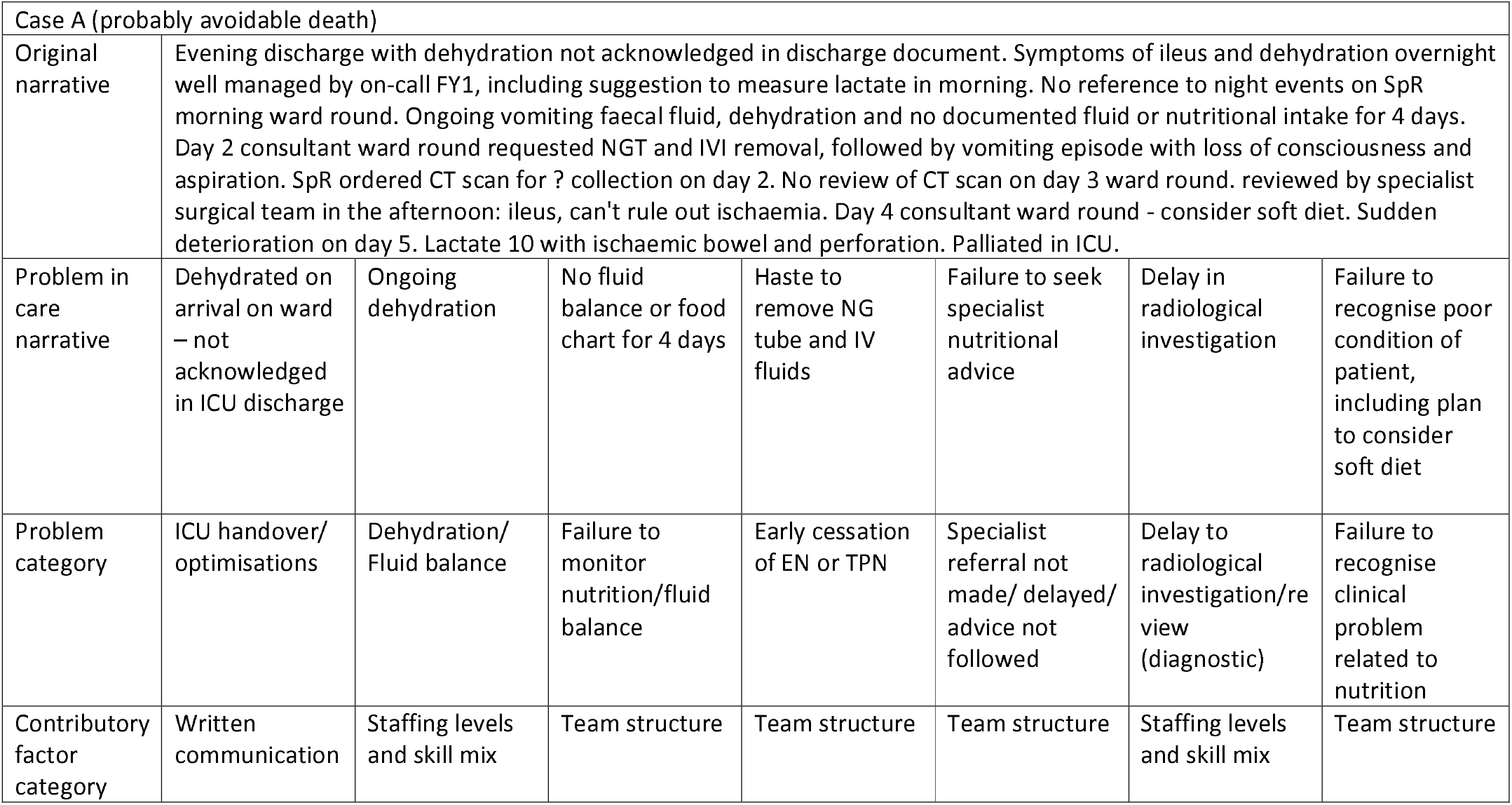
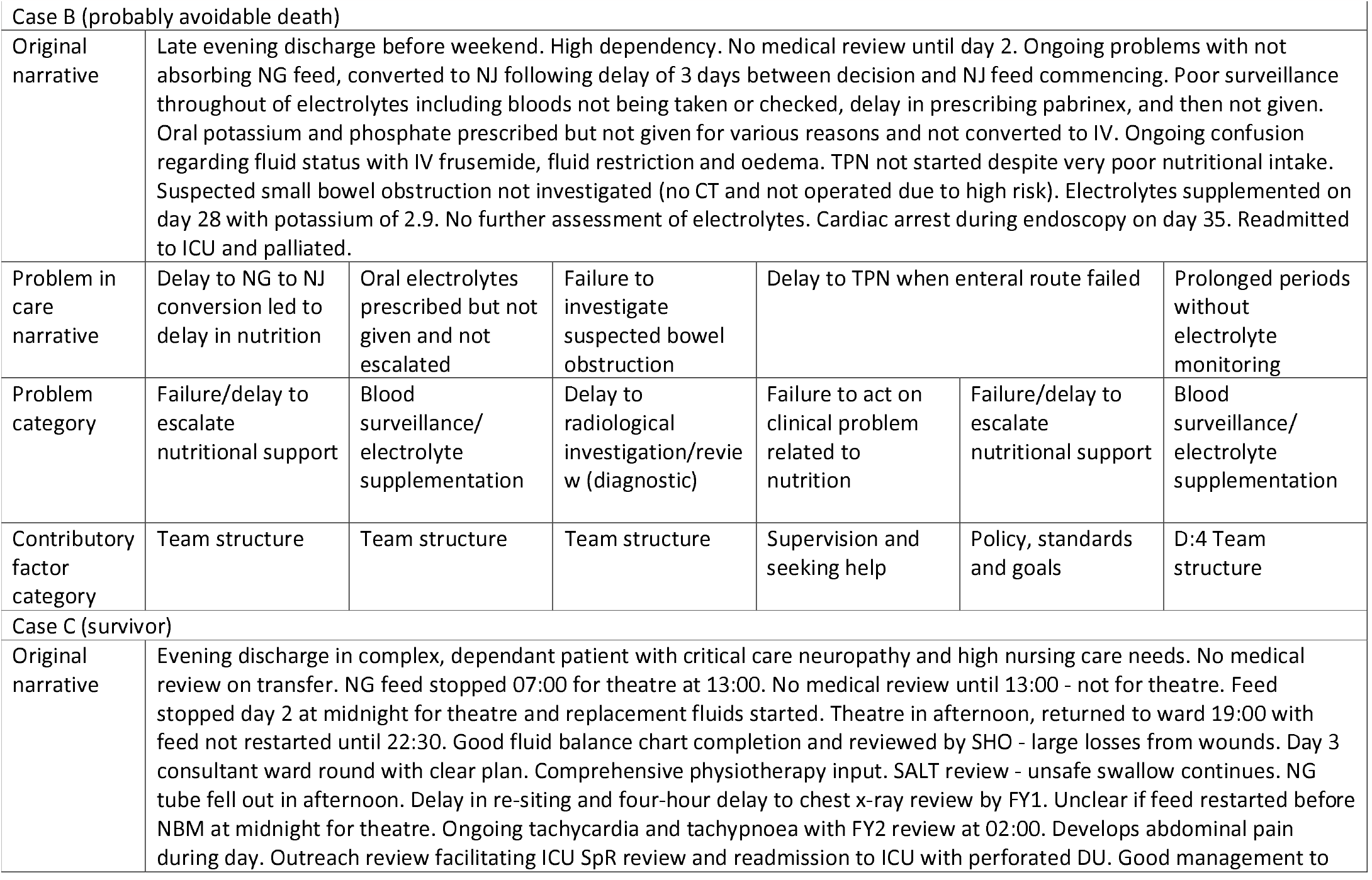
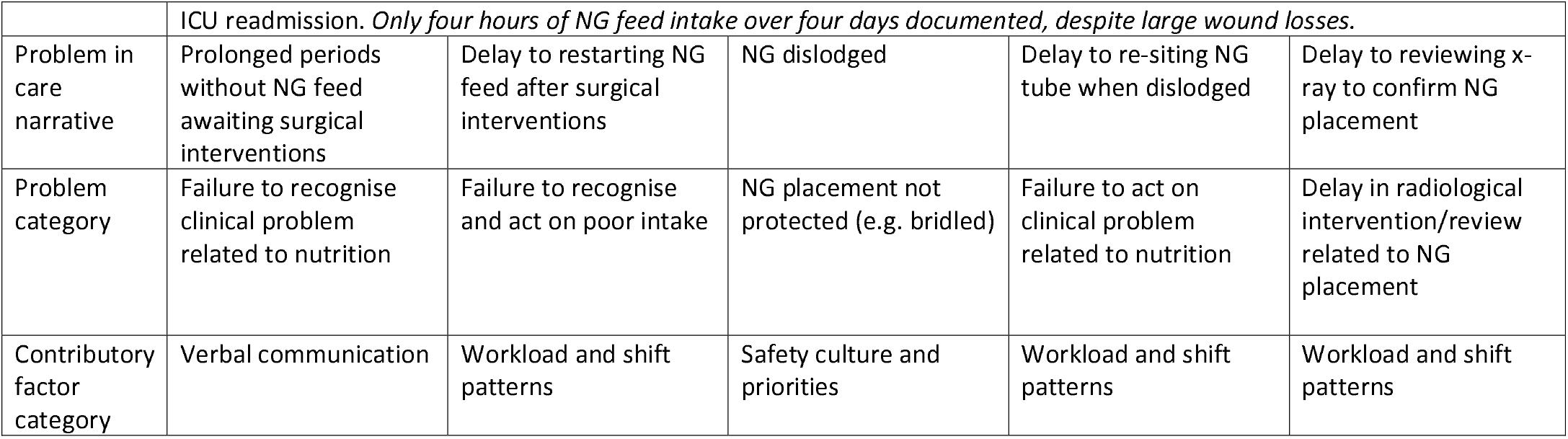
Illustrative vignettes

|  |  |  |  |  |  |  |  |
| --- | --- | --- | --- | --- | --- | --- | --- |
| Case A (probably avoidable death) |  |  |  |  |  |  |  |
| Original narrative | Evening discharge with dehydration not acknowledged in discharge document. Symptoms of ileus and dehydration overnight well managed by on-call FY1, including suggestion to measure lactate in morning. No reference to night events on SpR morning ward round. Ongoing vomiting faecal fluid, dehydration and no documented fluid or nutritional intake for 4 days. Day 2 consultant ward round requested NGT and IVI removal, followed by vomiting episode with loss of consciousness and aspiration. SpR ordered CT scan for ? collection on day 2. No review of CT scan on day 3 ward round. reviewed by specialist surgical team in the afternoon: ileus, can't rule out ischaemia. Day 4 consultant ward round - consider soft diet. Sudden deterioration on day 5. Lactate 10 with ischaemic bowel and perforation. Palliated in ICU. |  |  |  |  |  |  |
| Problem in care narrative | Dehydrated on arrival on ward – not acknowledged in ICU discharge | Ongoing dehydration | No fluid balance or food chart for 4 days | Haste to remove NG tube and IV fluids | Failure to seek specialist nutritional advice | Delay in radiological investigation | Failure to recognise poor condition of patient, including plan to consider soft diet |
| Problem category | ICU handover/optimisations | Dehydration/Fluid balance | Failure to monitor nutrition/fluid balance | Early cessation of EN or TPN | Specialist referral not made/ delayed/ advice not followed | Delay to radiological investigation/review (diagnostic) | Failure to recognise clinical problem related to nutrition |
| Contributory factor category | Written communication | Staffing levels and skill mix | Team structure | Team structure | Team structure | Staffing levels and skill mix | Team structure |

|  |  |  |  |  |  |  |
| --- | --- | --- | --- | --- | --- | --- |
| Case B (probably avoidable death) |  |  |  |  |  |  |
| Original narrative | Late evening discharge before weekend. High dependency. No medical review until day 2. Ongoing problems with not absorbing NG feed, converted to NJ following delay of 3 days between decision and NJ feed commencing. Poor surveillance throughout of electrolytes including bloods not being taken or checked, delay in prescribing pabrinex, and then not given. Oral potassium and phosphate prescribed but not given for various reasons and not converted to IV. Ongoing confusion regarding fluid status with IV frusemide, fluid restriction and oedema. TPN not started despite very poor nutritional intake. Suspected small bowel obstruction not investigated (no CT and not operated due to high risk). Electrolytes supplemented on day 28 with potassium of 2.9. No further assessment of electrolytes. Cardiac arrest during endoscopy on day 35. Readmitted to ICU and palliated. |  |  |  |  |  |
| Problem in care narrative | Delay to NG to NJ conversion led to delay in nutrition | Oral electrolytes prescribed but not given and not escalated | Failure to investigate suspected bowel obstruction | Delay to TPN when enteral route failed |  | Prolonged periods without electrolyte monitoring |
| Problem category | Failure/delay to escalate nutritional support | Blood surveillance/ electrolyte supplementation | Delay to radiological investigation/review (diagnostic) | Failure to act on clinical problem related to nutrition | Failure/delay to escalate nutritional support | Blood surveillance/ electrolyte supplementation |
| Contributory factor category | Team structure | Team structure | Team structure | Supervision and seeking help | Policy, standards and goals | D:4 Team structure |
| Case C (survivor) |  |  |  |  |  |  |
| Original narrative | Evening discharge in complex, dependant patient with critical care neuropathy and high nursing care needs. No medical review on transfer. NG feed stopped 07:00 for theatre at 13:00. No medical review until 13:00 - not for theatre. Feed stopped day 2 at midnight for theatre and replacement fluids started. Theatre in afternoon, returned to ward 19:00 with feed not restarted until 22:30. Good fluid balance chart completion and reviewed by SHO - large losses from wounds. Day 3 consultant ward round with clear plan. Comprehensive physiotherapy input. SALT review - unsafe swallow continues. NG tube fell out in afternoon. Delay in re-siting and four-hour delay to chest x-ray review by FY1. Unclear if feed restarted before NBM at midnight for theatre. Ongoing tachycardia and tachypnoea with FY2 review at 02:00. Develops abdominal pain during day. Outreach review facilitating ICU SpR review and readmission to ICU with perforated DU. Good management to |  |  |  |  |  |

|  |  |  |  |  |  |
| --- | --- | --- | --- | --- | --- |
|  | ICU readmission. *Only four hours of NG feed intake over four days documented, despite large wound losses.* |  |  |  |  |
| Problem in care narrative | Prolonged periods without NG feed awaiting surgical interventions | Delay to restarting NG feed after surgical interventions | NG dislodged | Delay to re-siting NG tube when dislodged | Delay to reviewing x-ray to confirm NG placement |
| Problem category | Failure to recognise clinical problem related to nutrition | Failure to recognise and act on poor intake | NG placement not protected (e.g. bridled) | Failure to act on clinical problem related to nutrition | Delay in radiological intervention/review related to NG placement |
| Contributory factor category | Verbal communication | Workload and shift patterns | Safety culture and priorities | Workload and shift patterns | Workload and shift patterns |

### Handover

We identified six instances where ICU handover of nutritional needs was poor or absent, including in vignette A, where the patient was dehydrated on ward arrival, without acknowledgement of this in the written ICU documentation.

### Nutritional monitoring and escalation

We identified failures in monitoring of nutritional intake or fluid balance in half of the cases (10/20) where nutritional problems were present (including vignette A). This impaired the ability of the multi-disciplinary team to recognise and act on poor nutritional intake (6/20 cases) and dehydration/fluid management problems (6/20 cases), preventing or delaying subsequent management.

We found ten cases where problems occurred with referrals to nutrition specialists (including dietetic, TPN and gastro-intestinal surgical teams), including failure or delay to refer, poor specialist advice being offered, and specialist advice not being followed. Problems with electrolyte imbalances (potassium, magnesium and phosphate) were commonly identified by specialist nutrition teams, and there were eight instances of failure to monitor or treat deranged electrolytes. Vignette B presents an extreme example of this with severe consequences for the patient.

Failing to recognise, escalate or act on nutrition problems was common, with 15 instances identified across the cases. Vignettes A and B illustrate the consequences of this, with slow recognition of small bowel obstruction/ileus having profound consequences for these patients. This was compounded by four cases of delay to undertake and/or review the reports of diagnostic radiological investigations, even when this had been discussed on ward rounds.

### Nutritional support provision

Where nutritional support was in place (usually delivered enterally), several problems with delivery were identified. In three cases, NG tubes were removed prior to establishing oral intake, including for vignette A, who was clearly documented as not able to manage oral intake. In four cases, the nasogastric (NG) tube was dislodged at least once, and only one patient had a bridled NG tube (a device securing the tube to the septum to prevent accidental removal). This, in combination with uncertainty about the NG tube being in the correct place, led to delays in resuming NG feed associated with the need to replace and/or confirm placement of the new tube by x-ray in two cases. In one case (vignette C) this process contributed to a prolonged failure to meet the nutritional needs of a patient with large exuding wounds. Lack of co-ordination of feeding and surgical interventions also contribute to this, resulting in only four hours of enteral feed documented as delivered over the course of four days.

We found four cases of delays in escalation of nutrition support where the current route had failed, leading to prolonged delays in meeting nutrition needs in each case. We identified four instances of poor team leadership, where poor nutritional delivery occurred over a prolonged period of time.

### Contributory human factors analysis

Due to the complexity, prolonged duration and multidisciplinary nature of nutritional support, the contributory human factor selected for most problems with nutrition was ‘team factors: team structure’ (56/81, table 4). Staffing levels and skill mix, and workload and shift patterns were both identified five times, and in four cases, individual competence was identified as contributing to a problem in care related to nutritional delivery. Table 2 includes the contributory human factors selected for each of the problems identified in the example vignettes (see supplementary material for a full list of the contributory factors categories).

**Table 4:**
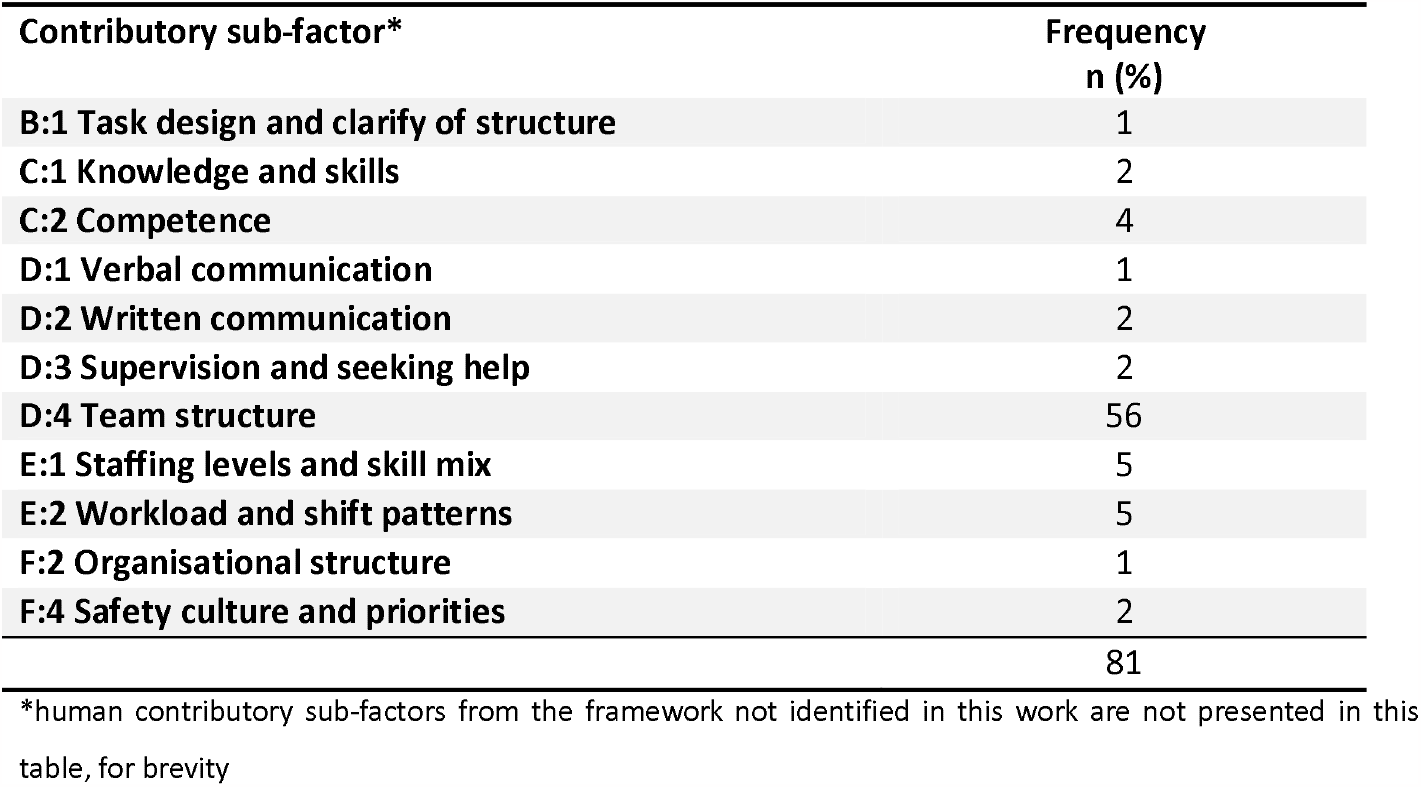
Contributory human factors to identified problems in care related to nutrition

| **Contributory sub-factor\*** | **Frequency**<br>**n (%)** |
| --- | --- |
| **B:1 Task design and clarify of structure** | 1 |
| **C:1 Knowledge and skills** | 2 |
| **C:2 Competence** | 4 |
| **D:1 Verbal communication** | 1 |
| **D:2 Written communication** | 2 |
| **D:3 Supervision and seeking help** | 2 |
| **D:4 Team structure** | 56 |
| **E:1 Staffing levels and skill mix** | 5 |
| **E:2 Workload and shift patterns** | 5 |
| **F:2 Organisational structure** | 1 |
| **F:4 Safety culture and priorities** | 2 |
|  | 81 |

\*human contributory sub-factors from the framework not identified in this work are not presented in this table, for brevity

### Semi-structured interviews

To develop our understanding of the context of post-ICU care delivery, we conducted interviews with a wide variety of stakeholders to gain multiple perspectives. Despite being fundamental to patient care and recovery, nutrition was not mentioned in any of the interviews with patients and family members, and only occasionally directly discussed by the 30 staff members interviewed. One foundation year doctor discussed a general reluctance to start TPN, identifying this as ‘false optimism’ that the patient would regain an enteral nutrition route, rather than ‘negligence’ in not providing nutrition (table 5, quote 1).

**Table 5:**
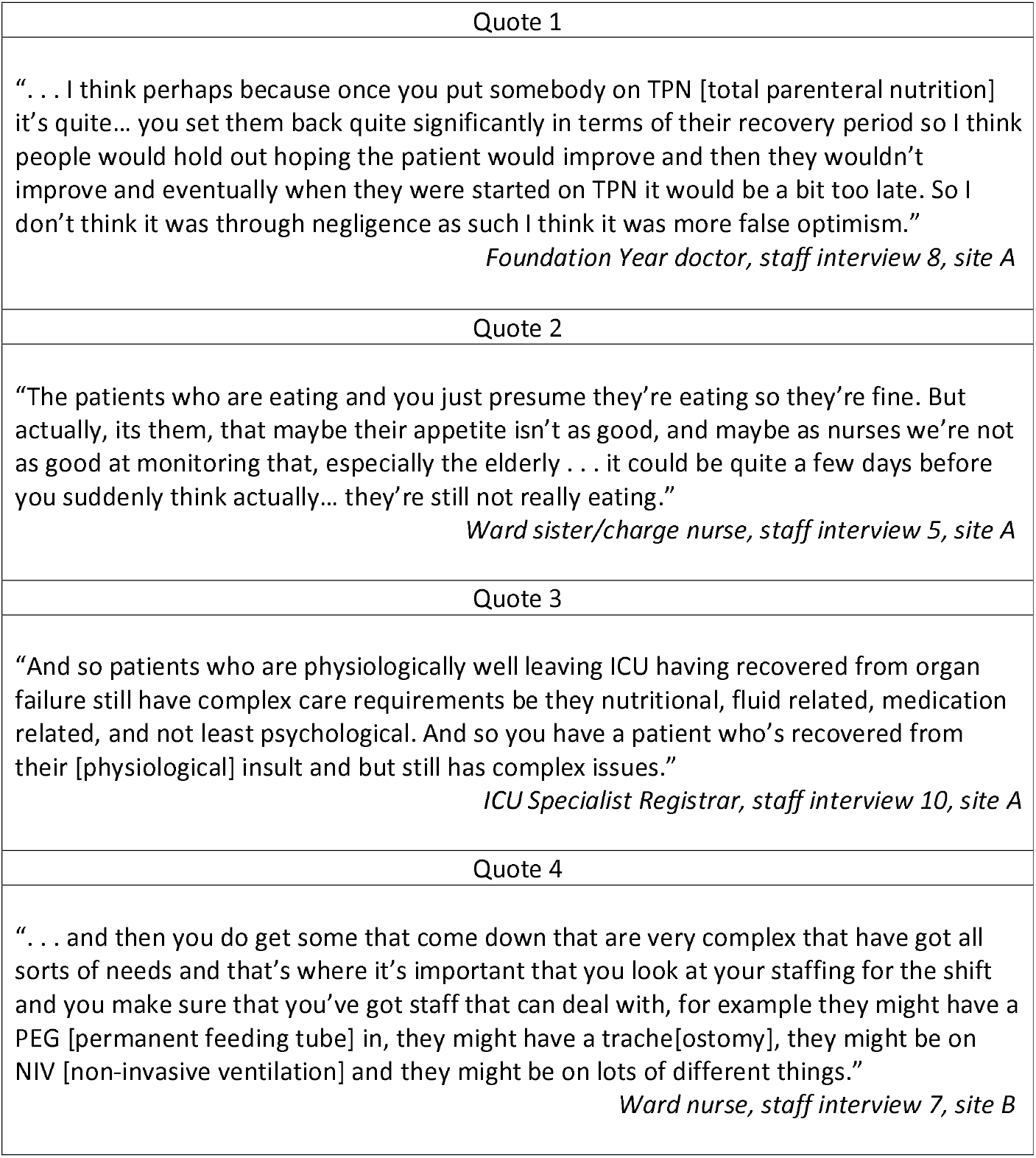
Illustrative interview quotes

| Quote 1 |
| --- |
| “ . . . I think perhaps because once you put somebody on TPN [total parenteral nutrition] it's quite... you set them back quite significantly in terms of their recovery period so I think people would hold out hoping the patient would improve and then they wouldn't improve and eventually when they were started on TPN it would be a bit too late. So I don't think it was through negligence as such I think it was more false optimism.”<br>*Foundation Year doctor, staff interview 8, site A* |
| Quote 2 |
| “The patients who are eating and you just presume they're eating so they're fine. But actually, its them, that maybe their appetite isn't as good, and maybe as nurses we're not as good at monitoring that, especially the elderly . . . it could be quite a few days before you suddenly think actually... they're still not really eating.”<br>*Ward sister/charge nurse, staff interview 5, site A* |
| Quote 3 |
| “And so patients who are physiologically well leaving ICU having recovered from organ failure still have complex care requirements be they nutritional, fluid related, medication related, and not least psychological. And so you have a patient who's recovered from their [physiological] insult and but still has complex issues.”<br>*ICU Specialist Registrar, staff interview 10, site A* |
| Quote 4 |
| “ . . . and then you do get some that come down that are very complex that have got all sorts of needs and that's where it's important that you look at your staffing for the shift and you make sure that you've got staff that can deal with, for example they might have a PEG [permanent feeding tube] in, they might have a trache[ostomy], they might be on NIV [non-invasive ventilation] and they might be on lots of different things.”<br>*Ward nurse, staff interview 7, site B* |

**Table 6:** Output of FRAM detailing the process of providing enteral nutrition to a patient discharged from ICU to the ward

| **Function 1: Recognising poor intake** |  |
| --- | --- |
| Input: | ICU handover (NG feeding required/established)<br>Nurse assessment<br>Ward round assessment |
| Pre-conditions: | Documentation of nutritional intake<br>Nutrition screening documented |
| Resources: | Nurse time |
| Controls: | Local policy on nutrition monitoring/screening |
| Time: | Prompt assessment to prevent prolonged period without nutrition |
| Output: | Dietician referral |
| **Function 2: Dietician assessment and feed prescription** |  |
| Input: | Dietician referral |
| Pre-conditions: | Documentation of nutritional intake<br>Blood results |
| Resources: | Nurse time<br>Dietician time and skill<br>Access to electronic notes system |
| Controls: | Local policy<br>NICE guidelines |
| Time: | Prompt assessment to prevent prolonged period without nutrition |
| Output: | Identified as needing support to maximise oral intake, or<br>Identified as needing NG feeding and feed prescribed |
| **Function 3: Clinical staff (re)insert new nasogastric tube** |  |
| Input: | Patient requires NG tube |
| Pre-conditions: | Patient consent<br>Patient condition appropriate for insertion |
| Resources: | Clinician time and skill<br>NG tube of correct type and size and associated equipment |
| Controls: | Local NG tube policy |
| Time: | n/a |
| Output: | NG tube in place |
| **Functions 3-7: Radiological confirmation of tube placement** |  |
| Input: | Requirement to start NG feeding |
| Pre-conditions: | NG tube in place |
| Resources: | Access to electronic patient record<br>Team/on-call doctor time to order chest x-ray<br>Radiology time to take x-ray<br>Porter time to take patient to department (optional)<br>Radiologist time and expertise to review and report on x-ray<br>Team/on-call doctor time to review x-ray report and confirm placement to nurse |
| Controls: | Local policy |
| Time: | Timeliness of feed administration to meet nutritional needs |
| Output: | NG tube placement in stomach confirmed<br>Tube incorrectly placed |
| **Function 8: Checking placement of nasogastric tube** |  |
| Input: | Requirement to (re)start NG feeding |
| Pre-conditions: | NG tube in place |
| Resources: | pH test and associated equipment<br>Nurse knowledge and skill<br>Nurse time |
| Controls: | Local policy |
| Time: | Timeliness of feed administration to meet nutritional needs |
| Output: | NG tube placement in stomach confirmed<br>NG tube placement not confirmed (tube dislodged/no aspirate obtained/aspirate pH test inconclusive) |
| **Function 9: Commence/continue feeding** |  |
| Input: | NG placement in stomach confirmed<br>NG feed prescription |
| Pre-conditions: | NG tube placement confirmed |
| Resources: | Prescribed feed<br>Feeding pump<br>Feeding giving set<br>Nurse knowledge |
| Controls: | Local policy |
| Time: | Timeliness of feed administration to meet nutritional needs |
| Output: | NG feed started |
| **Function 10: Pause feeding** |  |
| Input: | Need to pause feeding (break/NBM for procedure/placement concern) |
| Pre-conditions: | Communication between MDT of need to pause feeding and when<br>NG feed prescription (if prescribed feed break) |
|  | Visual check of NG tube placement (e.g. after mobilisation) |
| Resources: | Access to electronic prescription system<br>Nurse knowledge |
| Controls: | Local policy |
| Time: | Timeliness of feed pauses to reduce impact on nutritional intake |
| Output: | NG feed paused |

This was echoed by a ward sister who identified patients most at risk of malnutrition as those who were deemed able to eat, but whose intake was poor but not being routinely monitored (table 5, quote 2). However, although nutritional management was rarely directly discussed, staff frequently described the high care need of post-ICU patients, including nutritional support needs, as being challenging to manage on the ward (table 5, quotes 3 and 4).

### Functional Resonance Analysis – Enteral Feeding

Although the FRAM encompassed a wide range of conditions related to delivering nutrition to post-ICU patients on the ward, it is not possible to report all of these within the confines of this paper. Identification of poor intake and provision of enteral nutrition on the ward were identified as key areas for improvement in post-ICU nutrition from the in-depth reviews. We have therefore focused on reporting functions related to these issues. The full FRAM diagram is included in supplementary material (figure 1).

**Figure 1.**
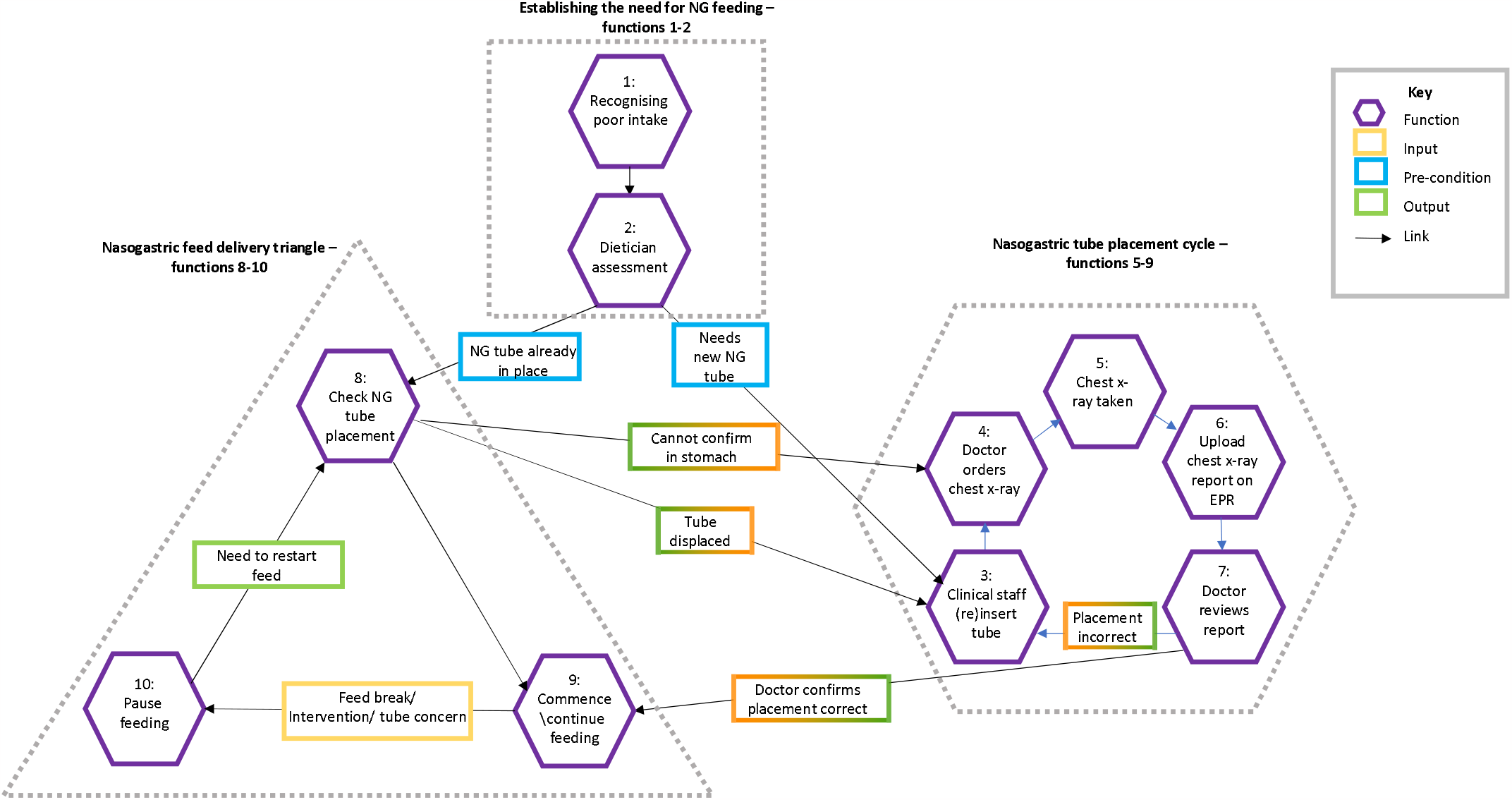
Simplified FRAM diagram of nasogastric feeding on the ward.

The process of providing enteral feeding on the ward is not linear. We identified 10 key functions essential to supporting adequate enteral nutrition provision on the ward., with three clusters of functions within this process: establishing the need for enteral feeding; the nasogastric tube placement cycle; and enteral feed delivery. Functions are summarised in figure 1, with all identified conditions presented in table 5. Outputs of one function often become inputs of the next (highlighted in the figure using two-tone boxes). We have summarised the process below.

### Establishing the need for enteral feeding

We identified two functions leading to prescription of enteral feeding (highlighted by the dotted square in figure 1). Function 1 – recognising a patient as having poor nutritional intake – had two possible inputs: enteral feeding may have already occurred in ICU and be part of the written handover, or poor intake may be recognised on the ward by nursing or medical staff. Adequate nutritional monitoring is a pre-condition for this function, contributing to the assessment of intake. This was absent in half of the in-depth reviews where nutritional problems were present, limiting the ability of staff to recognise poor nutrition intake. Where nutritional intake was recognised as poor, the output was dietician referral.

The output of function one – dietician referral – becomes the input of function two: dietician review. Resources for this function include dietician time and access to blood results. Pre-conditions include documentation of prior nutritional intake (already identified as commonly absent). Controls include local feeding guidelines. The common outputs of this function are to monitor and support oral intake, or to deliver nasogastric (NG) feeding.

### The nasogastric tube placement cycle

We identified a cycle of functions related to placing a nasogastric tube (dotted hexagon in figure 1). If not already in place, the clinician (usually a nurse but could be another professional with the required skills) inserts an NG tube (function 3). Resources for this function include clinician time, skills and equipment. Pre-conditions include appropriate patient condition and consent.

Once the tube has been sited, at all three sites the local protocol was for radiological confirmation of correct placement (a control). This requires several functions (4-7, dotted hexagon in figure 1) to occur in succession – a doctor must order the chest x-ray (4); the chest x-ray must be taken (5); the report from the x-ray must be uploaded into the electronic patient record (6); and the doctor must review the report (7). The main resource for each of the functions in this cycle is staff time. Where time is limited the process is delayed. Delays in each step of this process may cumulate into prolonged periods without nutrition. The doctor must then communicate one of two outputs to the result to nursing staff. If the output of this cycle is that the tube is incorrectly placed, this cycle returns to function 3: clinical staff (re)site tube, and the full process occurs again, incurring further delay. If the output is that the tube is correctly placed, this cycle ends and moves on to the next group of functions.

### Nasogastric feed delivery

We identified three key functions in this process (8-10, dotted triangle in figure 1). If the input is NG tube placement radiologically confirmed, the process can start at function 9 (commence feeding). If the input is NG tube already in place (and placement has not been radiologically confirmed), before feeding can be commenced/recommenced following a break, local policy at all three sites requires NG tube placement to be re-confirmed (function 8). Placement is confirmed by visual inspection of the tube markings at entry to the nostril against documentation, and pH testing of tube aspirate (confirming the aspirate as gastric contents). Resources for this function include staff time and expertise, and equipment to conduct the test. The control for this function is local policy. If placement is confirmed, the process can move on to Function 9. Alternative outputs include: NG tube dislodged or visibly moved; no aspirate obtained; and pH not within range. These outputs would require a return to the placement confirmation cycle, either at function 3 (clinical staff (re)insert tube) if the tube is dislodged, or function 4 (doctor orders chest x-ray) if the tube cannot be confirmed as in the stomach.

Once correct tube positioning is confirmed, nursing staff may commence or continue NG feeding (function 9). Pre-conditions for this function include a prescription. Resources include nurse time and skill, access to correct feed, and equipment to deliver the feed.

Once the feed is running, at some point it will be paused (function 10). Pre-conditions for pausing feed include prescribed feeding breaks, being nil by mouth for surgery or other interventions, or concerns about tube placement. Where planned surgical interventions are subsequently delayed or postponed to the following day, patients are at risk of prolonged periods without nutrition (time) (see Vignette C). The process returns to function 8 if/when NG feeding is planned to recommence.

This process is not linear. Although the first two functions occur in numerical order, the process will then either proceed to the nasogastric tube placement cycle if the is no NG tube already in place (at function 3), or to the NG feed delivery process (at function 8). At this point, if the NG tube cannot be confirmed as in the correct place at function 8, the process may still need to proceed to the placement cycle (at function 4), or continue in the delivery process. At any point in the delivery process, the tube may become dislodged or placement checks fail, at which point the process will return to the placement cycle – either at function 3 or 4 depending on the output of function 8 (tube displaced, or cannot confirm in stomach).

## DISCUSSION

Using a mixed methods approach has allowed us to investigate the underlying reasons and consequences of poor nutrition delivery for post-ICU patients. In-depth reviews allowed exploration of the implications of this failure of handover and ongoing nutrition provision, and vignettes drawn from these in-depth reviews helped illustrate the problems identified. Interviews with staff explored the context of care delivery and why these problems may have occurred. Finally, the FRAM detailed the structural process of enteral delivery in the ward environment.

Problems related to nutritional delivery were extensive, encompassing failure to monitor intake, failure to escalate to specialist input, haste in removing enteral feeding tubes before adequate oral intake was established, and poor management of enteral nutrition. These problems appeared to be due in part to a failure to appreciate the overall nutritional status of dependent patients rather than viewing their care day by day. In contrast, the better nutritional management identified in survivors may reflect their lower frailty compared with patients who died in hospital following their ICU stay, and therefore ability to manage their own nutritional needs.

### Multi-professional teamwork

The in-depth reviews demonstrated the wide variety of professionals involved in providing post-ICU nutritional support, including nurses, doctors, dieticians and specialist nutrition teams. This was reflected by the high frequency of ‘team structure’ identified as an underlying contributory human factor, demonstrating that good nutritional support relies on multi-disciplinary collaboration. This was also demonstrated by the FRAM, where multi-professional working was essential to ensure timely delivery of adequate enteral nutrition.

### Identifying malnutrition and monitoring nutritional intake

Identifying poor nutritional intake is a key input in the FRAM, delivered through various means, such as during ward rounds, nursing assessment or dietetic review. All these routes, however, relied on adequate monitoring of nutritional intake, to provide evidence of the need for nutritional support. Opportunities to refer to dietetic or other specialist support was identified as often missed in the in-depth reviews, as exemplified in vignette A. Staff interviews suggested this level of monitoring was often undeliverable within the limitations of ward-based workloads. Nutritional support, including monitoring, is a fundamental part of ward-based care, but does require a significant proportion of nursing time. Nurses receiving patients from ICU to the ward have described post-ICU patients as having higher care needs than they are able to meet within the ward workload [25,26]. In this context, nutrition delivery may not be prioritised, because the impact of not feeding a patient may not be as clearly apparent as the failure to deliver other therapies [27]. Lack of clarity in who is responsible for identifying and, especially, escalating poor oral intake (e.g. doctors, nurses or dieticians) may also contribute to this problem [27].

### Early removal of feeding tubes

Removal of feeding tubes before oral intake is established was also identified as a problem in care in the in-depth reviews, as exemplified by vignette A. This was suggested during interviews to be due to pressure to promote recovery, and a drive towards discharge. The FRAM identified the process of (re)placing enteral feeding tubes as at risk of prolonged delays, suggesting tubes should only be removed once it is clearly established they are no longer needed. Previous studies have also identified a cultural drive to remove feeding tubes to promote oral intake and rehabilitation [28,29]. However, previous research has shown that patients recovering from critical illness have greater nutritional deficits when receiving oral diet than enteral feeding [28,30,31]. Other studies found oral intake was limited by barriers such as nausea, vomiting and poor appetite following critical illness [31,32]. Although there are concerns that enteral feeding may supress appetite, this has been shown not be the case [33]. Avoiding early removal of NG tubes, and continuing combination feeding, may facilitate adequate nutrition until oral intake is fully established [9,11,34].

### Delivery of enteral nutrition

The in-depth reviews identified multiple issues with the process of delivering enteral nutrition. Vignette C describes the impact of this, with large gaps in enteral feeding due both to feed breaks for subsequently cancelled surgical interventions, and issues with nasogastric tube placement. The FRAM illustrates two processes where opportunities for significant delays in enteral feeding occur: the tube placement cycle and the feed delivery triangle (figure 1). For any patient recovering from critical illness, these cumulative delays may have a significant impact on their rehabilitation.

Previous studies have also demonstrated poor delivery of nutrition in post-ICU patients [30,31,35–37]. A key factor in poor enteral nutrition delivery was fasting for repeated procedures, resulting in prolonged breaks to planned feeding, similar to those described in vignette C [28,31]. Implementation of fasting guidelines may increase the percentage of prescribed feed delivered and reduce the hours of fasting but may be hindered by unpredictable procedure timings[7,38]. Although within-rather than post-ICU, these findings are similar to those in the in-depth reviews.

### Strengths and limitations

This study had several strengths. We published the protocol and registered our mixed methods project [17]. We collected data at three sites, selected to represent different sized hospitals and different post-ICU service provision. Our mixed methods approach to investigating post-ICU ward care allowed detailed exploration of common problems in care judged to have contributed to post-ICU in-hospital deaths. In addition to nutrition, we have similarly examined rehabilitation provision and out-of-hours discharge from ICU [19,20]. The FRAM brought rich mixed methods data from this project together, to map the process of delivering enteral nutrition in the ward environment, identifying some of the barriers which contributed to the poor nutritional support identified in the in-depth reviews.

As the REFLECT study focused on the post-ICU ward care generally, data related to nutrition were limited. The in-depth reviews relied on documentation [16], and we identified a common lack of nutritional monitoring documentation, which limited our ability to assess nutritional intake. The number of cases reviewed was also relatively small at 40. Despite this, we were able to identify a large number of problems in this area, with rich detail related to these problems. Indeed, given the reliance on documentation we may have under-estimated the magnitude of the problems which existed in post-ICU nutrition for our study population. Qualitative interview data were very limited in direct reference to nutrition provision, although there was significant discussion about the high workload associated with post-ICU patients more generally. Rather than a limitation, this does highlight the lack of importance placed on nutritional support by both patients and staff.

Categorisation of the nutrition problems was expanded from the established framework, with code developed from the narratives. These codes are subjective and may have been differently categorised by a different reviewer. However, that the original framework only included one category for nutrition: 13: Other – nutrition, demonstrates the lack of importance placed on nutrition provision within clinical care.

### Implications for practice and future research

Our mixed methods approach has given us insight into the structural complexity of delivering nutrition to patients discharged from ICU to a ward setting. By using this approach, we are able to make several recommendations which may contribute to the goal recommended by Heyland et al of delivering 80-90% of prescribed nutritional requirements for critical care survivors, beyond ICU discharge [39]. These are: close nutrition monitoring on the ward; early referral to specialist nutrition teams including dietetics; avoidance of early NG tube removal (before oral intake established); and prioritisation of the steps in the NG tube insertion process, to ensure timely and adequate enteral nutrition.

Despite identifying that research is required into the role of nutrition in the post-acute period, there remains little evidence related to this [40]. Bear et al. call for studies investigating nutrition and exercise together, into the post-ICU recovery period [41].

Finally, although our work focused on post-ICU patients, our findings are likely to be transferrable to other word-based cohorts who are physically dependent and/or have high care needs.

## CONCLUSION

Nutrition delivery was identified as a significant problem contributing to avoidable deaths in the previously published RCRR, with absence of a clear nutritional plan on ICU discharge common [18]. Using a novel mixed methods approach has allowed us to explore the consequences and underlying reasons for the significant issues in post-ICU nutritional support we identified. Common problems included failure to monitor intake, delay to refer to nutritional specialists, and haste to remove nasogastric feeding tubes. Staff identified that these problems occurred due to limitations in the system of care delivery on the ward. The FRAM identified multiple points in the process of delivering enteral nutrition where significant delays could be incurred. Addressing these problems requires collaborative multi-professional working to ensure patients receive the nutrition they required to maximise their recovery from critical illness.

## Data Availability

All data produced in the present study are available upon reasonable request to the authors

## Acknowledgements

The authors gratefully acknowledge the patients, family members and staff who participated in the REFLECT study, and the sites who supported us in the conduct of this research.

## REFERENCES

[1] Rawal G, Yadav S, Kumar R. Post-intensive care syndrome: An overview. J Transl Int Me. 2017;5:90–2. https://doi.org/10.1515/jtim-2016-0016.

[2] Inoue S, Hatakeyama J, Kondo Y, Hifumi T, Sakuramoto H, Kawasaki T, et al. Post-intensive care syndrome: its pathophysiology, prevention, and future directions. Acute Medicine & Surgery 2019;6:ams2.415. https://doi.org/10.1002/ams2.415.

[3] Bein T, Bienvenu OJ, Hopkins RO. Focus on long-term cognitive, psychological and physical impairments after critical illness. Intensive Care Me. 2019;45:1466–8. https://doi.org/10.1007/s00134-019-05718-7.

[4] Gerth AMJ, Hatch RA, Young JD, Watkinson PJ. Changes in health-related quality of life after discharge from an intensive care unit: a systematic review. Anaesthesia 2019;74:100–8. https://doi.org/10.1111/ANAE.14444.

[5] Puthucheary ZA, Rawal J, McPhail M, Connolly B, Ratnayake G, Chan P, et al. Acute Skeletal Muscle Wasting in Critical Illness Original Investigation | CARING FOR THE CRITICALLY ILL PATIENT. JAMA 2013;310:1591–600. https://doi.org/10.1001/jama.2013.278481.

[6] Moisey LL, Merriweather JL, Drover JW. The role of nutrition rehabilitation in the recovery of survivors of critical illness: underrecognized and underappreciated. Crit Car. 2022;26:1–17. https://doi.org/10.1186/S13054-022-04143-5.

[7] Singer P, Reintam Blaser A, Berger MM, Alhazzani W, Calder PC, Casaer MP, et al. ESPEN Guideline ESPEN guideline on clinical nutrition in the intensive care unit. Clinical Nutritio. 2019;38:48–79. https://doi.org/10.1016/j.clnu.2018.08.037.

[8] Whitehead J, Summers MJ, Louis R, Weinel LM, Lange K, Dunn B, et al. Assessment of physiological barriers to nutrition following critical illness. Clinical Nutritio. 2022;41:11–20. https://doi.org/10.1016/J.CLNU.2021.11.001.

[9] van Zanten ARH, de Waele E, Wischmeyer PE. Nutrition therapy and critical illness: Practical guidance for the icu, post-icu, and long-term convalescence phases. Crit Car. 2019;23:1–10. https://doi.org/10.1186/S13054-019-2657-5/FIGURES/2.

[10] Bear DE, Wandrag L, Merriweather JL, Connolly B, Hart N, Grocott MPW. The role of nutritional support in the physical and functional recovery of critically ill patients: A narrative review. Crit Car. 2017;21:1–10. https://doi.org/10.1186/S13054-017-1810-2/TABLES/1.

[11] Lambell KJ, Tatucu-Babet OA, Chapple LA, Gantner D, Ridley EJ. Nutrition therapy in critical illness: a review of the literature for clinicians. Crit Car. 2020;24. https://doi.org/10.1186/S13054-020-2739-4.

[12] Wischmeyer PE. Nutrition Therapy in Sepsis. Crit Care Cli. 2018;34:107–25. https://doi.org/10.1016/j.ccc.2017.08.008.

[13] Merriweather JL, Salisbury L, Walsh T, Smith P. Nutritional care after critical illness: A qualitative study of patients’ experiences. Journal of Human Nutrition and Dietetic. 2016;29:127–36. https://doi.org/10.1111/jhn.12287.

[14] Elm E von, Altman DG, Egger M, Pocock SJ, Gøtzsche PC, Vandenbroucke JP. Strengthening the Reporting of Observational Studies in Epidemiology (STROBE) statement: guidelines for reporting observational studies. BMJ 2007;335:806–8. https://doi.org/10.1136/BMJ.39335.541782.AD.

[15] Tong A, Sainsbury P, Craig J. Consolidated criteria for reporting qualitative research (COREQ): a 32-item checklist for interviews and focus groups. International Journal for Quality in Health Car. 2007;19:349–57. https://doi.org/10.1093/INTQHC/MZM042.

[16] Hutchinson A. Using the structured judgement review method A guide for reviewers. London: 2017.

[17] Vollam S, Gustafson O, Hinton L, Morgan L, Pattison N, Thomas H, et al. Protocol for a mixed-methods exploratory investigation of care following intensive care discharge: The REFLECT study. BMJ Ope. 2019;9. https://doi.org/10.1136/bmjopen-2018-027838.

[18] Vollam S, Gustafson O, Young JD, Attwood B, Keating L, Watkinson P. Problems in care and avoidability of death after discharge from intensive care: a multi-centre retrospective case record review study. Crit Car. 2021;25:10. https://doi.org/10.1186/s13054-020-03420-5.

[19] Vollam S, Gustafson O, Morgan L, Pattison N, Thomas H, Watkinson P. Patient Harm and Institutional Avoidability of Out-of-Hours Discharge From Intensive Care: An Analysis Using Mixed Methods. Crit Care Me. 2022;50:1083–92. https://doi.org/10.1097/CCM.0000000000005514.

[20] Gustafson OD, Vollam S, Morgan L, Watkinson P. A human factors analysis of missed mobilisation after discharge from intensive care: A competition for care? Physiotherapy 2021;113:131–7. https://doi.org/10.1016/j.physio.2021.03.013.

[21] Hogan H, Healey F, Neale G, Thomson R, Black N, Vincent C. Learning from preventable deaths: exploring case record reviewers’ narratives using change analysis. J R Soc Me. 2014;107:365–75. https://doi.org/10.1177/0141076814532394.

[22] Braun V, Clarke V. Using thematic analysis in psychology. Qual Res Psycho. 2006;3:77– 101. https://doi.org/10.1191/1478088706qp063oa.

[23] Hollnagel E. FRAM: The Functional Resonance Analysis Method. CRC Press; 2017. https://doi.org/10.1201/9781315255071.

[24] Clay-williams R, Hounsgaard J, Hollnagel E. Where the rubber meets the road1: using FRAM to align work-as-imagined with work-as-done when implementing clinical guidelines. Implementation Scienc. 2015;10:1–8. https://doi.org/10.1186/s13012-015-0317-y.

[25] Enger R, Andershed B. Nurses’ experience of the transfer of ICU patients to general wards: A great responsibility and a huge challenge. J Clin Nur. 2018;27:e186–94. https://doi.org/10.1111/jocn.13911.

[26] Kauppi W, Proos M, Olausson S. Ward nurses’ experiences of the discharge process between intensive care unit and general ward. Nurs Crit Care 2018:1–7. https://doi.org/10.1111/nicc.12336.

[27] Chapple LS, Chapman M, Shalit N, Udy A, Deane A, Williams L. Barriers to Nutrition Intervention for Patients With a Traumatic Brain Injury: Views and Attitudes of Medical and Nursing Practitioners in the Acute Care Setting. Journal of Parenteral and Enteral Nutritio. 2018;42:318–26. https://doi.org/10.1177/0148607116687498.

[28] Chapple LS, Chapman M, Shalit N, Udy A, Deane A, Williams L. Barriers to Nutrition Intervention for Patients With a Traumatic Brain Injury: Views and Attitudes of Medical and Nursing Practitioners in the Acute Care Setting. Journal of Parenteral and Enteral Nutritio. 2018;42:318–26. https://doi.org/10.1177/0148607116687498.

[29] Merriweather JL, Smith P, Walsh T. Nutritional rehabilitation after ICU-does it happen: A qualitative interview and observational study. J Clin Nur. 2014;23:654–62. https://doi.org/10.1111/jocn.12241.

[30] Ridley EJ, Parke RL, Davies AR, Bailey M, Hodgson C, Deane AM, et al. What happens to nutrition intake in the post-intensive care unit hospitalization period? An observational cohort study in critically ill adults. Journal of Parenteral and Enteral Nutritio. 2019;43:88–95. https://doi.org/10.1002/jpen.1196.

[31] Chapple L anne S, Deane AM, Heyland DK, Lange K, Kranz AJ, Williams LT, et al. Energy and protein deficits throughout hospitalization in patients admitted with a traumatic brain injury. Clinical Nutritio. 2016;35:1315–22. https://doi.org/10.1016/J.CLNU.2016.02.009.

[32] Merriweather JL, Griffith DM, Walsh TS. Appetite during the recovery phase of critical illness: a cohort study. European Journal of Clinical Nutrition 2018 72:7 2018;72:986– 92. https://doi.org/10.1038/s41430-018-0181-3.

[33] Stratton RJ, Stubbs RJ, Elia M. Short-Term Continuous Enteral Tube Feeding Schedules Did Not Suppress Appetite and Food Intake in Healthy Men in a Placebo-Controlled Trial. J Nut. 2003;133:2570–6. https://doi.org/10.1093/JN/133.8.2570.

[34] Bear DE, Wandrag L, Merriweather JL, Connolly B, Hart N, Grocott MPW. The role of nutritional support in the physical and functional recovery of critically ill patients: A narrative review. Crit Car. 2017;21:1–10. https://doi.org/10.1186/S13054-017-1810-2/TABLES/1.

[35] Jarden RJ, Sutton-Smith L, Boulton C. Oral intake evaluation in patients following critical illness: an ICU cohort study. Nurs Crit Car. 2018;23:179–85. https://doi.org/10.1111/NICC.12343.

[36] Moisey LL, Pikul J, Keller H, Yeung CYE, Rahman A, Heyland DK, et al. Adequacy of Protein and Energy Intake in Critically Ill Adults Following Liberation From Mechanical Ventilation Is Dependent on Route of Nutrition Delivery. Nutrition in Clinical Practic. 2021;36:201–12. https://doi.org/10.1002/NCP.10558.

[37] Wittholz K, Fetterplace K, Clode M, George ES, MacIsaac CM, Judson R, et al. Measuring nutrition-related outcomes in a cohort of multi-trauma patients following intensive care unit discharge. Journal of Human Nutrition and Dietetic. 2020;33:414– 22. https://doi.org/10.1111/JHN.12719.

[38] Jenkins B, Calder PC, Marino L V. Evaluation of implementation of fasting guidelines for enterally fed critical care patients. Clinical Nutritio. 2019;38:252–7. https://doi.org/10.1016/J.CLNU.2018.01.024/ATTACHMENT/8EE8C963-7AAC-456C-B891-FA67B57ECA83/MMC1.PDF.

[39] Heyland DK, Cahill N, Day AG. Optimal amount of calories for critically ill patients: Depends on how you slice the cake! Crit Care Med 2011;39:2619–26. https://doi.org/10.1097/CCM.0B013E318226641D.

[40] Latronico N, Herridge M, Hopkins RO, Angus D, Hart N, Hermans G, et al. The ICM research agenda on intensive care unit-acquired weakness. Intensive Care Me. 2017;43:1270–81. https://doi.org/10.1007/S00134-017-4757-5/FIGURES/3.

[41] Bear DE, Griffith D, Puthucheary ZA. Emerging outcome measures for nutrition trials in the critically ill. Curr Opin Clin Nutr Metab Car. 2018;21:417–22. https://doi.org/10.1097/MCO.0000000000000507.

